# Reanalysis of the Pfizer mRNA BNT162b2 SARS-CoV-2 vaccine data fails to find any increased efficacy following the boost: Implications for vaccination policy and our understanding of the mode of action

**DOI:** 10.1101/2021.02.23.21252315

**Authors:** Allan Saul, Heidi E. Drummer, Nick Scott, Tim Spelman, Brendan S. Crabb, Margaret Hellard

## Abstract

**Background:** In clinical trials two vaccinations with mRNA vaccines have shown high efficacy in preventing COVID-19. However, in the context of a pandemic, the time to generation of protective immunity, the need for and timing of a second vaccination are matters of legitimate debate. This manuscript explores the efficacy and timing of the second dose COVID-19 vaccines, including a reanalysis of data from the Pfizer mRNA BNT162b2 mRNA SARS-CoV-2 vaccine phase 3 study.

**Methods and findings:** A non-weighted three-segment, two knot linear regression was fitted to the published cumulative infection incidence from the Pfizer BNT162b2 vaccine Phase III trial using the lspine routine in R. The optimal knot days were estimated through sensitivity analysis and the confidence limits for efficacy estimates were determined by Monte Carlo Simulations. This analysis showed the vaccine was effective from day 11 post first vaccination. The estimated efficacy over the period 11 to 28 days post first vaccination was 0.94 and there was no detectable increase in efficacy following the second vaccination. The efficacy post first vaccination substantially preceded the development of detectable serum neutralizing antibody.

**Conclusions:** Strongly protective immunity develops rapidly following a single vaccination and at least in the short period covered by the timetable of the Phase III trial, there was no additional benefit from a second vaccination. This increases options for use of this vaccine, e.g., for ring fence vaccination, for use in travelers and for mass vaccination rollout. It highlights the need for further research into duration of immunity following a single vaccination and for understanding mechanisms of protection.

## Introduction

The current COVID-19 pandemic has made the development and deployment of vaccine a high priority and the Research, Pharmaceutical, Regulatory and Medical communities have responded in developing and deploying vaccines in a remarkably short time. At the time of writing, several vaccines have been licensed for use, including two vaccines based on mRNA technology: Pfizer BNT162b2 vaccine[1] and the Modena mRNA-1273 SARS-CoV-2 vaccine[2] that are the subject of this paper. Thus far, the mRNA vaccines receiving authorization of the basis of Phase 3 studies require two vaccinations, either 3 or 4 weeks apart.

Several vaccines are now deployed but in the face of still tragic deaths rates, there is major pressure for rapid deployment. There remain serious practical issues with the rate at which the vaccines can be manufactured and people vaccinated. To address this issue, one response has been to give as many people as possible a single vaccination and to delay the second vaccination. This raises the question of the efficacy of a single immunization.

As judged by protection against infection by SARS-CoV-2, measured by PCR positive nasal samples, in studies in Israel, Chodick et al estimated an efficacy of 51% comparing infection rates on days 13 to 21 with rates from day 0 to 12[3] but noted that the infection rates in vaccinees deviating from about day 19 following vaccination from the infection rates in the same subjects immediately post-vaccination. In a subsequent re-analysis of these data, Hunter and Brainard[4] concluded that the single vaccination gives about 90% protection against infection by 21 days post first vaccination and then levelled off.

As judged by protection against COVID-19 disease, the data showing the infection rates in vaccinees versus the control subjects are remarkable for both the Pfizer and Moderna vaccines: As expected for both vaccines, the infection rates in the vaccinees followed the infection rates in the controls for the first few days then showed an impressive decrease approximately two weeks after the first vaccination. However, interestingly neither showed any obvious further decrease in the already low infection rates following the second vaccination following day 21 (Pfizer) or day 28 (Moderna). Specifically, there was no downward inflection in the slope of the graphs of percentage infected vs time at any time following the second vaccination.

There is a second striking aspect of the data reported from both the Pfizer BNT162b2 vaccine and Modena mRNA-1273 SARS-CoV-2 vaccine: at the time of the second vaccination, at least half the subjects had serum neutralizing antibody titers that were undetectable or below the lower limit of quantification and the remainder, although positive, had relatively low titers. Despite a large increase in serum neutralization titers following the second vaccination, there was no corresponding increase in apparent efficacy of the vaccine. Since serum neutralization titers have been used as a guide for developing vaccines and for assessing the potential of vaccines to protect against new variants, this result is unexpected.

In this note, the Pfizer Phase 3 data is re-examined, and we show that over the initial time period not only is there substantial protection from a single vaccination but there is no apparent additional protection from the second vaccination. Since this protection also significantly precedes the development of neutralizing antibody in the serum, this observation also raises substantial questions about both the correlates and the mechanisms of protection in the period immediately following the first vaccination.

## Methods

### Data

The cumulative % of people infected as a function of time in the vaccinated and control groups were derived from Figure 3 of the BNT162b2 mRNA vaccine phase III trial report, for both vaccinated and placebo groups from day 1 to day 111 following the first vaccination [1]. Similarly, cumulative % were also derived from Figure 3 in the Modena mRNA-1273 SARS-CoV-2 vaccine trial report. However, in the Moderna study, the number of cases of COVID-19 in the control and vaccine groups were too low over the first few weeks to allow detailed analysis and the placebo group had a significant non-linear increase. Therefore, they were not used for a detailed analysis but subsequently used for a comparison with the Pfizer results.

### Regression modelling

A linear regression of cumulative percent infected in the placebo group as a function of time gave a good fit to the data (intercept −0.094 s.e. 0.014%; slope 0.0204 s.e. 0.00021 % per day; Adjusted R^2^ 0.988). Therefore, we assume that the cumulative percentage of vaccinees infected would also be linear, with potentially different slopes for three distinct time periods: the initial period following first vaccination (with the same theoretical slope as the placebo group, i.e. the time period prior to the development of immunity); the period from when protective immunity was developed following the first vaccination to when further protective immunity was developed following the second vaccination; and the remaining period to the end of the surveillance.

We fitted a three segment non-weighted linear regression to the cumulative % infected data using the lspline function in R[5] with two knots corresponding to the days at which the immunity was postulated to develop following the first and second immunizations, respectively. We determined the most appropriate knot days by testing the fit of a range of models, as judged by the adjusted R^2^, where the first knot varied between day 5 to day 15 and the second knot varied between day 22 to day 32. The best fit was always with a first knot on day 11 but varying the second knot made little difference (Adjusted R2 varied from 0.9866 to 0.9867) so for subsequent analyses the first knot was set at day 11 and the second at day 28, the day that immune responses were measured following the second vaccination.[6] Code for the sensitivity analysis is listed in the Supplement.

Vaccine efficacies for the period Day 11 to Day 28 and for Day 28 to Day 111 were calculated as

E_11-28_ = (1-(Slope day 11 to day 28)/(slope day 0 to day 11)) and

E_28-111_ = (1-(Slope day 28 to day 111)/(slope day 0 to day 11)), respectively.

Where E_11-28_ is the efficacy in the vaccine group over day 11 to 28 and E_28-111_ is the efficacy over day 28 to 111.

As expected from a time series, an analysis of the regression and its residual showed substantial heteroskedasticity, autocorrelation between the observed cumulative distribution and non-normal distribution of residuals. Therefore, estimates of confidence intervals for the estimated efficacies and power of the analysis were done by simulation.

Confidence intervals for efficacy estimates were determined with a purpose written R routine using 5000 Monte Carlo simulations of the data set, assuming 18560 subjects were present throughout the time-period (the number of people receiving two vaccinations and who were followed for a median of 2 months). For each of the 5000 simulations, a Poisson random number generator was used to assign the number of cases each day, with mean derived from the value of the fitted three-segment linear model.

### Power

A power calculation was performed by Monte Carlo simulation to estimate the minimum difference in efficacy following the first vaccination (day 11 to day 28) compared to following the second vaccination (day 28 to day 111) that could be detected, given the trial size, infection rates and duration of each time period. For a hypothetical E_11-28_ ranging from 0.5 to 1.0 and an E_28-111_ of 0.94, we calculated the proportion of 5000 simulations where the 2.5% lower limit in the simulated range of E_28-111_ was greater than E_11-28_.

Codes for all procedures are listed in the Supplement.

## Results

### Pfizer BNT162b2 mRNA vaccine

The optimal day for the first knot as judged by the adjusted R^2^ was always day 11 (Fig. 1), independent of the day of the second knot. Varying the second knot made little difference; for example, with a first knot on day 11, the Adjusted R^2^ varied between 0.9866 to 0.9867 when the second knot varied from day 21 to day 32.Therefore knot 1 on day 11 and knot 2 on day 28 were used for subsequent analyses. Day 28 corresponds to the day on which post vaccine 2 immunological analysis was conducted.[1]

**Figure 1.**
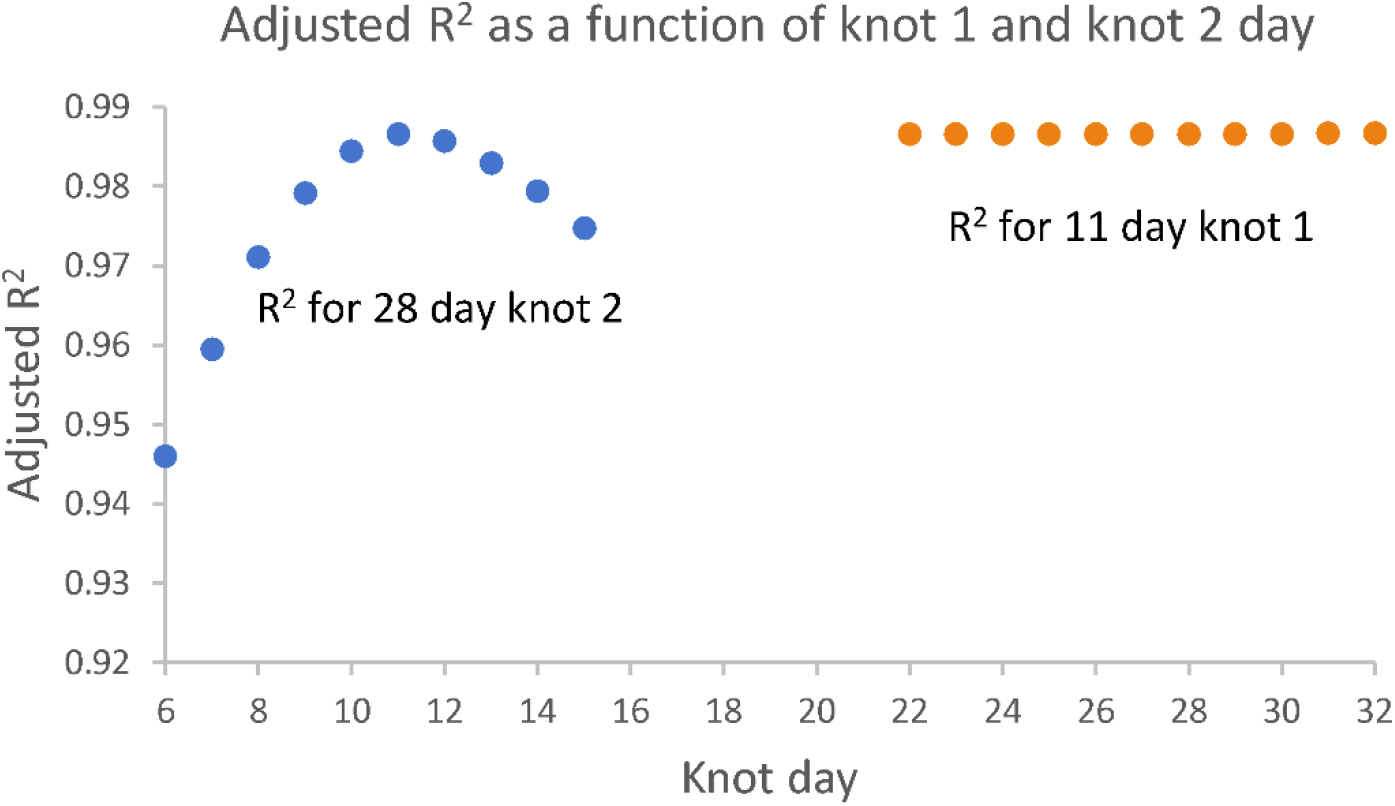
**Adjusted R**^**2**^ **obtained by fitting a three-segment linear model to the observed cumulative % infected vaccinated subjects as a function of the day for the first knot (blue), assuming a 28-day second knot and for the second knot (orange) assuming an 11 -day first knot**.

The fit to the observed Phase III trial data was obtained with day 11 as the first knot and day 28 as the second knot is shown in Figure 2.

**Figure 2.**
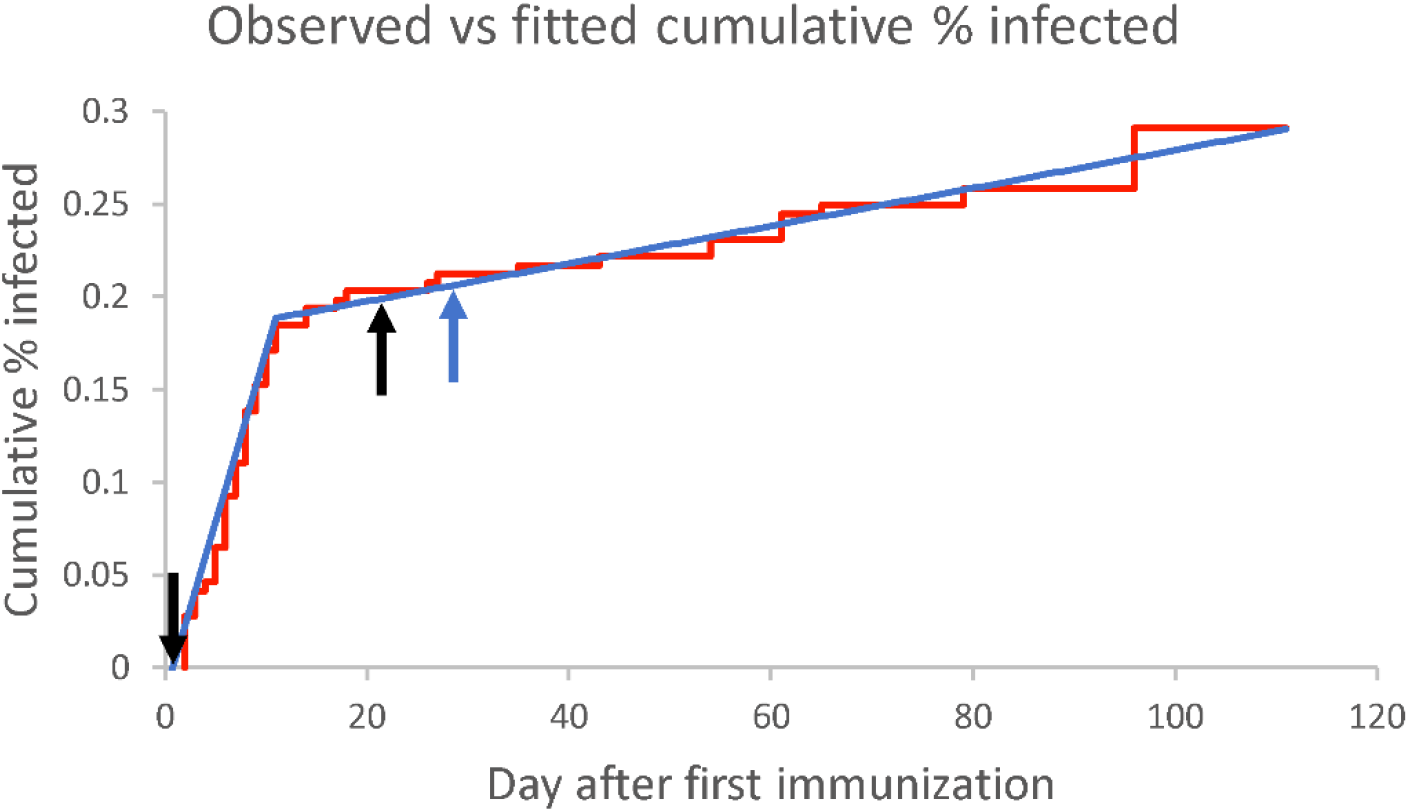
**Observed cumulative % of vaccinees infected (data from the Pfizer et al paper[1], Figure 3) is shown in red with the fitted three linear segment model shown in blue. Black arrows indicate the vaccination days. The blue arrow (day 28) is the joint between the second and third fitted linear segments and is the day where an inflection would indicate an increase in protection following the second vaccination.**

The slopes of the three segments of the linear model for the percentage of the vaccine group infected vs time, as well as the estimated slope for the initial 11 days of the placebo group, are shown in the first column of Table 1. The second column of Table 1 shows the corresponding estimates of efficacy, with confidence intervals determined by Monte Carlo simulations of the data set. Intercepts from regression analysis for both placebo day 1-11 and vaccine day 1 to 11 were not significantly different to zero.

**Table 1.** **Estimates and standard errors for the linear fits of the % infection vs day post first vaccination for placebo group and vaccine groups**.

| Vaccine & Time | Slope* (95% CI)<sup>†</sup> | Efficacy (95% CI)<sup>†</sup> |
| --- | --- | --- |
| Placebo D1 to D11 | 0.0195 |  |
| Vaccine D1 to D11 | 0.0191 (0.0137 to 0.0271) |  |
| Vaccine D11 to D28 | 0.0000940 (-0.00054 to 0.0028) | 0.951 (0.843 to 1.026) |
| Vaccine D28 to D111 | 0.0001022 (0.00056 to 0.0017) | 0.946 (0.904 to 0.974) |

\*estimated by linear regression on the reported data
<sup>†</sup>estimated by simulation analysis

The estimated efficacy from the simulation analysis from day 11 to day 28 and after day 28 are not significantly different to the published efficacy (0.948) from day 29 to day 111. The mean ratio of E_11-28_ to E_28 −111_ was 1.005 (95% CI 0.898 to 1.098).

For E_11-28_ ranging from a hypothetical 0.5 to 1, the percentage of 5000 simulations where E_11-28_ was less than the 2.5% lower limit of the simulated range for the efficacy from the day 28 to day 111 simulations is shown in Figure 3. If E_28-111_ is 0.94, there is an 80% power of observing E_11-28_ < E_28-111_ if the hypothetical E_11-28_ was <0.86, or a 95% power if the hypothetical E_11-28_ was < 0.8

**Figure 3.**
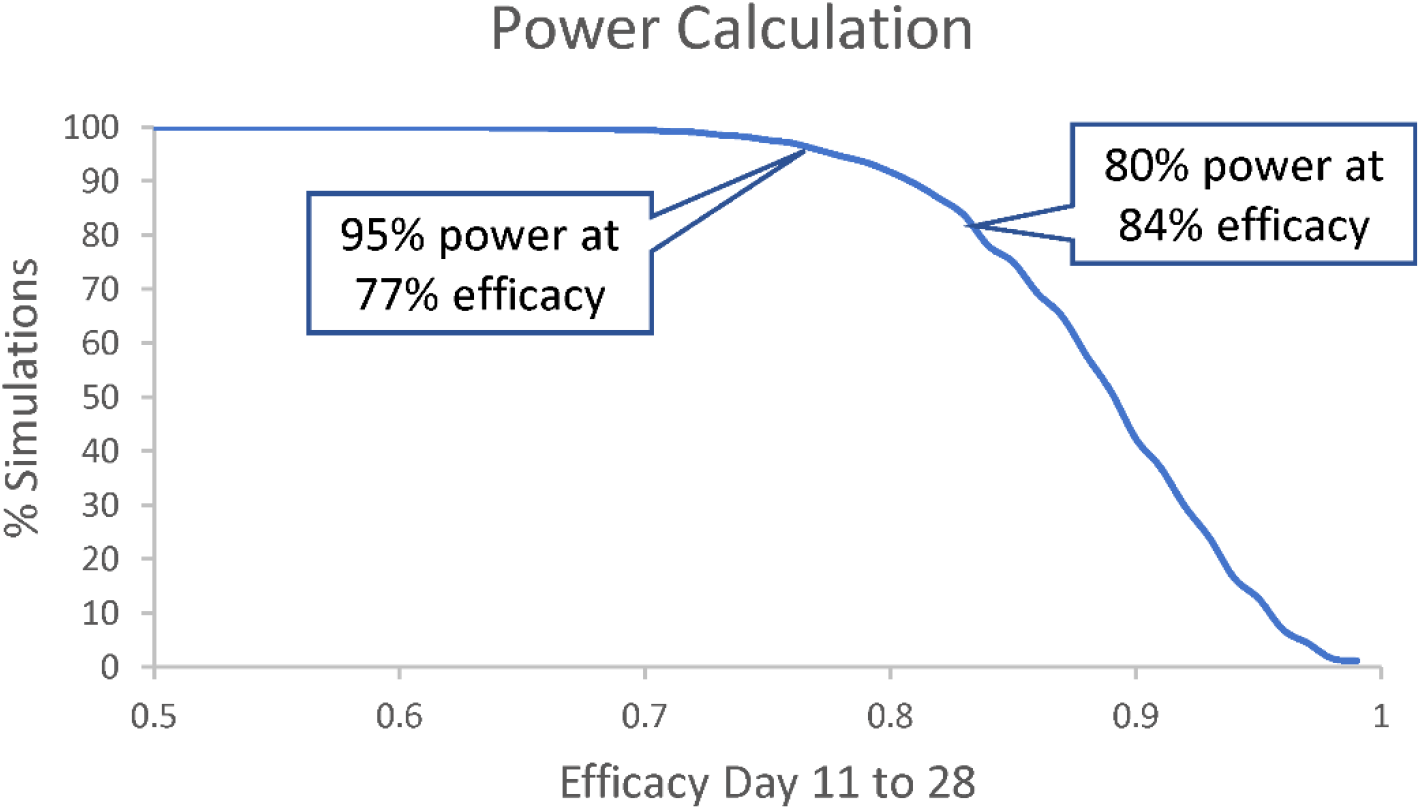
**Power to detect a difference in efficacy before or after the second vaccination. Graph shows the proportion of simulations using a range of hypothetical E**_**11-28**_ **from 0**.**5 to 1 that gave a calculated efficacy of E**_**11-28**_ **that were below the 2**.**5% lower limit of the range of simulations from a hypothetical E**_**28-111**_ **of 0**.**94**

### Modena mRNA-1273 SARS-CoV-2 vaccine

The second vaccination for the Moderna vaccine was on day 28, so the assumed knot day for change in efficacy after the second vaccine was day 35 (i.e., 28 + 7 days). There were insufficient cases in the period from first vaccination to day 11 to calculate efficacy using the spline fit methods and a significant curvature in the % placebo group infected with time also complicated a regression analysis. Therefore, the E_11-35_ and E_35-118_ were calculated using the number of cases in the vaccine groups compared to the placebo groups over these time periods. Note that these time periods differ from the time periods used in the Moderna trial analysis. The calculated E_11-35_ and E_35-118_ were 0.87 (95% CI 0.56 to 0.96) and 0.94 (95% CI 0.89 to 0.97), respectively. These are not significantly different.

## Discussion

These results are surprising: the Pfizer BNT162b2 mRNA vaccine appears to achieve full efficacy approximately 11 days following a single vaccination with no observable increase in efficacy following the second vaccination. The efficacy estimates calculated using the infection rates in the vaccinees from day 11 to day 28 and day 28 to day 111, compared to the infection rates in the vaccinees from day 1 to day 11, are indistinguishable at 0.95. The calculated efficacy rates are similar to the efficacy calculated in the Phase III trial by comparing infection rates in vaccine and placebo groups from day 28 on.

The power simulation suggest that these estimates are robust. Had the E_11-28_ efficacy been substantially lower, it would have been detected (80% power of detecting an efficacy lower than 0.86 at p=0.025, and a 95% power of detecting an efficacy of less than 0.80).

Although the power to detect a difference is much lower, the results with the Moderna mRNA-1273 SARS-CoV-2 vaccine are consistent with the results from the Pfizer vaccine: no significant difference in the measured efficacy from 11 to 35 days after the first vaccine and from 7 days after the second vaccination (i.e., day 35) (Efficacy 0.87 vs 0.94, respectively).

Not only is the lack of observed difference in the efficacy rate after the first and second vaccine unexpected, but they also raise major questions about the underlying mechanism of protection elicited by these vaccines. For the Pfizer BNT162b2 mRNA vaccine Phase 1 study, the amount of IgG binding to the S1 domain and the neutralization titers were measured in volunteers at 21 days and 28 days. There was approximately a 10-fold increase in the GMT for antibody binding between day 21 and day 28[7].

Considerable emphasis has been put on the levels of neutralizing antibody in the serum of vaccinated animals and humans for predicting efficacy of a vaccine[8] and ability to protect against new variants[9]. In the case of both the Pfizer and Moderna vaccines, the levels of neutralizing antibody were low prior to the second vaccination. In the results from the Phase I study, only four of the twelve 18-55 year-old subjects vaccinated with 30 µg Pfizer vaccine (i.e., the dose use in the efficacy studies) had measurable neutralizing antibody titers in serum [7]. The large increase in neutralizing titer from a geometric mean of 14 to 361 seven days following the second vaccination does not appear to be accompanied by an increase in efficacy. Similarly, in the Phase 1 study of the Moderna vaccine, only eight of the 15 subjects receiving the 100 µg dose has measurable serum neutralizing antibody on the day of the second vaccination with a geometric mean titer of 18 raising to 256 seven days following the second vaccination[6, 9]. Although the observed efficacy against nasal carriage was slower to develop in the Israeli study than the efficacy against disease, the protection against carriage also precedes the full development of serum antibody capable of binding to the spike or neutralizing virus.

These results are cause for optimism. Efficacy against disease with the mRNA starts at about 11 days. This opens up possibilities where rapid short-term protection that could be used for “ring fencing” vaccination following an outbreak or cluster. Also, it could be given to travelers in the fortnight prior to their planned trip. Given there was no demonstrated immediate improvement in giving a second vaccination after 3 or 4 weeks it also raises the potential to provide a single dose (at least in the short term when supplies are limited) and provide health services with more flexibility in administering vaccines and in the timing of a second dose if found to be warranted.

These results raise important research questions that require urgent exploration:

- How long does protection last after a single vaccination with the mRNA vaccines? Given the decreasing incidence of COVID-19 in many countries [10] and the roll-out of licensed vaccines, the ethics of doing a vaccine trial to compare single versus two doses and a planned trial would be complicated. However, there will inevitably be people vaccinated with the Pfizer or Moderna vaccine that fail to get a second vaccination at the scheduled time. Monitoring the infection rates in these people must be a high priority.
- What are the mechanisms of protection against disease and against carriage [3] following a single vaccination with mRNA vaccines? Several types of immune responses in vaccinees have not been reported that may play a role in protection following vaccination or natural infection. For example, mRNA vaccines can be potent inducers of innate immunity [11, 12]. SARS-CoV-2 specific secreted dIgA, has been proposed as important following natural infection [13]. Antibody effector functions may also contribute to protection [14, 15]. A better understanding of potential multifactorial protection mechanisms other than serum antibody binding or serum neutralizing antibody may be important for optimizing vaccine schedules, assessing the potential impact of new antigenic variants and developing new generation?anti-COVID-19 vaccines.

## Supporting information

R programs and data set

## Data Availability

Primary data is available from publications listed in the paper. Data derived from these publications are listed in the text and in a supplementary file

## Acknowledgments

None

## Abbreviations

E_11-28_, E_28-111_: Vaccine efficacies from day 11 to day 28 and from day 28 to day 111, respectively
s.e.: standard error
CI: confidence interval

## Supplementary Information

R routines for

- knot day sensitivity: mRNA_Knot_Sensitivity.R
- Calculating efficacy and confidence intervals by simulation: mRNA_Efficacy_CI.R
- power by simulation: mRNA_Efficacy_Power.R

Cumulative % infection in vaccine group from Fig. 3, Phase III trial data: PfizerC.txt

## References

1. Polack FP, Thomas SJ, Kitchin N, Absalon J, Gurtman A, Lockhart S, et al. Safety and Efficacy of the BNT162b2 mRNA Covid-19 Vaccine. N Engl J Med. 2020;383(27):2603–15. Epub 2020/12/11. doi: 10.1056/NEJMoa2034577. PubMed PMID : 33301246; PubMed Central PMCID: PMCPMC7745181.

2. Baden LR, El Sahly HM, Essink B, Kotloff K, Frey S, Novak R, et al. Efficacy and Safety of the mRNA-1273 SARS-CoV-2 Vaccine. N Engl J Med. 2021;384(5):403–16. Epub 2020/12/31. doi: 10.1056/NEJMoa2035389. PubMed PMID : 33378609; PubMed Central PMCID: PMCPMC7787219.

3. Chodick G, Tene L, Patalon T, Gazit S, Ben Tov A, D. Dc, et al. The effectiveness of the first dose of BNT162b2 vaccine in reducing the SARFS-CoV-2 infection 13-24 days after immunization: real world 2021 [cited 2021 February 23]. Available from: https://www.medrxiv.org/content/10.1101/2021.01.27.21250612v1.

4. Hunter PR, Brianard J. Estimating the effectiveness of the Pfizer COVID-19 BNT162b2 vaccine after a single dose. A reanalysis of a study of ‘real-world’ vaccination outcomes from Israel 2021 [cited 2021 February 23]. Available from: https://www.medrxiv.org/content/10.1101/2021.02.01.21250957v1.

5. Bojanowski M. lspline: Linear Splines with Convenient Parametrisations 2017 Feburary 23, 2021. Available from: https://cran.r-project.org/web/packages/lspline/lspline.pdf.

6. Jackson LA, Roberts PC, Graham BS. A SARS-CoV-2 mRNA Vaccine - Preliminary Report. Reply. N Engl J Med. 2020;383(12):1191–2. Epub 2020/08/20. doi: 10.1056/NEJMc2026616. PubMed PMID : 32813942.

7. Walsh EE, Frenck RW, Jr., Falsey AR, Kitchin N, Absalon J, Gurtman A, et al. Safety and Immunogenicity of Two RNA-Based Covid-19 Vaccine Candidates. N Engl J Med. 2020;383(25):2439–50. Epub 2020/10/15. doi:10.1056/NEJMoa2027906. PubMed PMID : 33053279; PubMed Central PMCID: PMCPMC7583697.

8. Sui Y, Bekele Y, Berzofsky JA. Potential SARS-CoV-2 Immune Correlates of Protection in Infection and Vaccine Immunization. Pathogens. 2021;10(2). Epub 2021/02/13. doi:10.3390/pathogens10020138. PubMed PMID : 33573221.

9. Weisblum Y, Schmidt F, Zhang F, DaSilva J, Poston D, Lorenzi JCC, et al. Escape from neutralizing antibodies by SARS-CoV-2 spike protein variants. bioRxiv. 2020. Epub 2020/08/04. doi:10.1101/2020.07.21.214759. PubMed PMID : 32743579; PubMed Central PMCID: PMCPMC7386497.

10. World Health Organization. https://www.who.int/publications/m/item/weekly-epidemiological-update 16-february-2021 2021 [cited 2021 February 20]. Available from: https://www.who.int/publications/m/item/weekly-epidemiological-update---16-february-2021.

11. Liang F, Lindgren G, Lin A, Thompson EA, Ols S, Rohss J, et al. Efficient Targeting and Activation of Antigen-Presenting Cells In Vivo after Modified mRNA Vaccine Administration in Rhesus Macaques. Mol Ther. 2017;25(12):2635–47. Epub 2017/09/30. doi:10.1016/j.ymthe.2017.08.006. PubMed PMID : 28958578; PubMed Central PMCID: PMCPMC5768558.

12. Wang Y, Zhang Z, Luo J, Han X, Wei Y. mRNA vaccine: a potential therapeutic strategy. Molecular Cancer. 2021;20. Epub February 16. doi:10.1186/s12943-021-01311-z.

13. Wang Z, Lorenzi JCC, Muecksch F, Finkin S, Viant C, Gaebler C, et al. Enhanced SARS-CoV-2 neutralization by dimeric IgA. Sci Transl Med. 2021;13(577). Epub 2020/12/09. doi:10.1126/scitranslmed.abf1555. PubMed PMID : 33288661; PubMed Central PMCID: PMCPMC7857415.

14. Hansen J, Baum A, Pascal KE, Russo V, Giordano S, Wloga E, et al. Studies in humanized mice and convalescent humans yield a SARS-CoV-2 antibody cocktail. Science. 2020;369(6506):1010–4. Epub 2020/06/17. doi:10.1126/science.abd0827. PubMed PMID : 32540901; PubMed Central PMCID: PMCPMC7299284.

15. Schafer A, Muecksch F, Lorenzi JCC, Leist SR, Cipolla M, Bournazos S, et al. Antibody potency, effector function and combinations in protection from SARS-CoV-2 infection in vivo. bioRxiv. 2020. Epub 2020/10/01. doi:10.1101/2020.09.15.298067. PubMed PMID : 32995782; PubMed Central PMCID: PMCPMC7523108.

