## Supplementary material for "Reanalysis of the Pfizer mRNA BNT162b2 SARS-CoV-2 vaccine data fails to find any increased efficacy following the boost: Implications for vaccination policy and our understanding of the mode of action": R programs and data set: mRNA_Efficacy_CI.R

```
#Program for calculating the efficacy from the data and to calculate confidence limits by simulation

library(lspline)

setwd(dirname(rstudioapi::getActiveDocumentContext()$path)) 
filepath<- getwd()

#data from Pfizer Phase 3 trial paper
CumCase<-scan(paste(filepath, "/PfizerC.txt", sep = ""))  #list of the % vaccinees infected on each of the 111 days following first vaccination 
MaxDay<-111
Day<-c(1:MaxDay)
k1<-11  # knot day 1
k2<-28  # knot day 2

#perform the 2 knot fit to the actual data and output the result

VFit.sum<-summary(lm(CumCase~lspline(Day,c(k1,k2),marginal=FALSE)))
capture.output(VFit.sum, file = paste(filepath,"/VFit_sum.txt",sep=""))


No.Placebo<-18530
No.Vaccine<-18556
PercentInfPlacebo<-2.254
EfficacyV1<-(1-VFit.sum$coefficients[3,1]/VFit.sum$coefficients[2,1])  # V1 indicates the period the immune response is manifest after first vaccination and before immunity is boosted with the second vaccination
EfficacyV2<-(1-VFit.sum$coefficients[4,1]/VFit.sum$coefficients[2,1])  # V2 indicates the period of the boosted immune response following the second vaccination

#program follows

PP<-PercentInfPlacebo*No.Placebo/(100*MaxDay) # ave number of people infected per day in placebo
PV1<-PP*(1-EfficacyV1) # ave expected to be infected per day in V1 period
PV2<-PP*(1-EfficacyV2) # ave expected to be infected per day in V2 period

Iter<-5000  #maximum 5000 to do a shapiro.test

VFit.out<-data.frame(matrix(0, ncol=7, nrow=Iter))
colnames(VFit.out)<-c("Intercept","S1","S2","S3","E1","E2","E2/E3")
VFit.CI<-data.frame(matrix(0,ncol=7,nrow=6))
colnames(VFit.CI)<-c("Intercept","S1","S2","S3","E1","E2","E2/E3")
rownames(VFit.CI)<-c("95% UL","90 UL","median","mean","90% LL","95%LL")

#calculate the distribution of the ratios of the simulated efficacy D11-28 to simulated efficacy D29-112 assuming they are really equal
# aim is to determine the 95% confidence intervals in the distribution of simulated ratio of efficacy

for (i in 1:Iter) {
  DayCase<-c(rpois(1:k1, lambda = PP),rpois((k1+1):k2, lambda = PV1),rpois((k2+1):MaxDay, lambda = PV2)) 
  CumCase<-cumsum(DayCase)*100/No.Vaccine
  VFit.sum<-summary(lm(CumCase~lspline(Day,c(11,28),marginal=FALSE)))
  VFit.out[i,1]<-VFit.sum$coefficients[1,1]
  VFit.out[i,2]<-VFit.sum$coefficients[2,1]  
  VFit.out[i,3]<-VFit.sum$coefficients[3,1]
  VFit.out[i,4]<-VFit.sum$coefficients[4,1]
  VFit.out[i,5]<-1-VFit.out[i,3]/VFit.out[i,2]
  VFit.out[i,6]<-1-VFit.out[i,4]/VFit.out[i,2]  
  VFit.out[i,7]<-VFit.out[i,5]/VFit.out[i,6] 
}  

for (i in 1:7){
#  VFit.out[order(VFit.out[,i])]
  SortD<-sort(VFit.out[,i])
  VFit.CI[1,i]<-SortD[0.025*Iter]
  VFit.CI[2,i]<-SortD[0.05*Iter] 
  VFit.CI[3,i]<-SortD[0.5*Iter]
  VFit.CI[4,i]<-mean(VFit.out[,i])  
  VFit.CI[5,i]<-SortD[0.95*Iter]
  VFit.CI[6,i]<-SortD[0.975*Iter]  
}

write.csv(VFit.CI,paste(filepath, "/VFit_CI.csv",sep=""))
```
