## Supplementary material for "Reanalysis of the Pfizer mRNA BNT162b2 SARS-CoV-2 vaccine data fails to find any increased efficacy following the boost: Implications for vaccination policy and our understanding of the mode of action": R programs and data set: mRNA_Efficacy_Power.R

```
#Program for calculating power to detect a difference in E11-28 and E28-111 by simulation

library(lspline)
setwd(dirname(rstudioapi::getActiveDocumentContext()$path)) 
filepath<- getwd()

#data from Pfizer Phase 3 trial paper
CumCase<-scan(paste(filepath, "/PfizerC.txt", sep = ""))  #list of the % vaccinees infected on each of the 111 days following first vaccination 
MaxDay<-111
Day<-c(1:MaxDay)

No.Placebo<-18530
No.Vaccine<-18556
PercentInfPlacebo<-2.254

#program follows

PP<-PercentInfPlacebo*No.Placebo/(100*MaxDay) # ave number of people infected per day in placebo
# PV1<-PP*(1-EfficacyV1) # average expected to be infected per day in V1 period
# PV2<-PP*(1-EfficacyV2) # average expected to be infected per day in V2 period

Iter<-5000  #maximum 5000 to do a shapiro.test

VFit.out<-data.frame(matrix(0, ncol=7, nrow=Iter))
colnames(VFit.out)<-c("Intercept","S1","S2","S3","E1","E2","E2/E3")


EfficacyV1<-0.49
EfficacyV2<-0.94

VFit.Power<-data.frame(matrix(0, ncol=2, nrow=50))
colnames(VFit.Power)<-c("EfficacyV1","% Power")

for (j in 1:50){
  EfficacyV1<-EfficacyV1+0.01 
  
  PV1<-PP*(1-EfficacyV1) # average expected to be infected per day in V1 period
  PV2<-PP*(1-EfficacyV2) # average expected to be infected per day in V2 period
  
  for (i in 1:Iter) {
    DayCase<-100*(c(rpois(1:11, lambda = PP),rpois(12:28, lambda = PV1),rpois(29:MaxDay, lambda = PV2)))/No.Vaccine
    CumCase<-cumsum(DayCase)
    VFit.sum<-summary(lm(CumCase~lspline(Day,c(11,28),marginal=FALSE)))
    

    
    VFit.out[i,1:4]<-as.numeric(VFit.sum$coefficients[1:4,1])
    VFit.out[i,5]<-1-VFit.out[i,3]/VFit.out[i,2]
    VFit.out[i,6]<-1-VFit.out[i,4]/VFit.out[i,2]
    VFit.out[i,7]<-VFit.out[i,6]/VFit.out[i,7]
    
  }

  SortV2<-sort(VFit.out[,6])
  LLV2<-SortV2[Iter*0.025]
  SortV1<-sort(VFit.out[,5])  
  
  VFit.Power[j,2]<-100*(which.min(abs(SortV1-LLV2)))/Iter
  VFit.Power[j,1]<-EfficacyV1
  
}
write.csv(VFit.Power,paste(filepath, "/VFit_Power_P.csv",sep=""))
```
