## Supplementary material for "Reanalysis of the Pfizer mRNA BNT162b2 SARS-CoV-2 vaccine data fails to find any increased efficacy following the boost: Implications for vaccination policy and our understanding of the mode of action": R programs and data set: mRNA_Knot_Sensivity.R

```
#Program for sensitivity analysis to find the optimal knot day for transistion following first vaccination
library(lspline)

setwd(dirname(rstudioapi::getActiveDocumentContext()$path))
filePath<- getwd()

CumCase<-scan(paste(filePath, "/PfizerC.txt", sep = ""))  #list of the % vaccinees infected on each of the 111 dcays following first vaccination
Day<-c(1:111)

MaxR2<-data.frame(matrix(0, ncol=3, nrow=121)) #Data frame storing the adjusted R^2 as a function of knot day
colnames(MaxR2)<-c("k1","k2","AdjR2")


for (k2 in 22:32) {
  for (k1 in 5:15) {
    MaxR2[k1-4+11*(k2-22),1]<-k1
    MaxR2[k1-4+11*(k2-22),2]<-k2
    MaxR2[k1-4+11*(k2-22),3]<-summary(lm(CumCase~lspline(Day,c(k1,k2),marginal=TRUE)))$adj.r.squared
  }
}  

Mk1<-MaxR2[which.max(MaxR2[,3]),1]
Maxk1<-subset(MaxR2, MaxR2$k2==28)
Maxk2<-subset(MaxR2, MaxR2$k1==Mk1)

write.csv(MaxR2,file=paste(filePath,"/MaxR2.csv",sep = ""))
write.csv(Maxk1,file=paste(filePath,"/Maxk1.csv",sep = ""))
write.csv(Maxk2,file=paste(filePath,"/Maxk2.csv",sep = ""))
```
